# A Parent-Generated Framework of Early Connection: Findings from a CBPR Qualitative Study

**DOI:** 10.64898/2026.06.12.26355487

**Authors:** Katrina Fuller, Sophia Duby, Diana More, Maeve Winter, Marissa Lanoff, Nicole Loveless, Jeannette Mejias, Desarae Smalls, Tiffany Solomon, Deepa Srinivasavaradan, Steven Thibert, Carly Vargas, Nikki Shearman, Dani Dumitriu, Andréane Lavallée

**Affiliations:** Center for Early Relational Health, Columbia University Irving Medical Center, New York, NY; Family Network Collaborative, Nurture Connection, Georgetown University, Washington, DC; Statewide Parent Advocacy Network, Newark, NJ; Children’s Hospital of Philadelphia, Philadelphia, PA; THRIVE Family Health & Education Center, Durham, NC; Reach Out and Read, Inc, Boston, MA; Department of Psychiatry, Columbia University Irving Medical Center, New York, NY

## Abstract

**Background:** Early relational health (ERH) constructs are derived from research observations rather than lived experiences. This study foregrounds diverse parent voices to examine how they describe connection with their young children.

**Methods:** Using community-based participatory research (CBPR), this study was co-designed with parent leaders from Reach Out and Read. A semi-structured interview guide was co-designed, and parent leaders subsequently conducted and transcribed 18 interviews with parents from their networks. Researchers analyzed transcripts using Reflexive Thematic Analysis. Member checking sessions with parent leaders informed the analytic framework.

**Results:** Six organizing principles were identified. (1) Parent-child connection begins with an instinctual sense of responsibility. (2) Connection ebbs and flows as parent and child adapt to one another through daily activities. (3) Family circumstances, including family structure, cultural expectations, and intergenerational values, directly shape this connection. (4) Parents’ own upbringings and past relationships indirectly shape how they connect with their child. (5) For connection to grow, parents must show up physically and emotionally for their children despite competing demands. (6) Parents grow through engaged parenting, and that growth feeds back into the connection, creating a self-sustaining cycle of relational health.

**Conclusions:** Our analysis generated two constructs underspecified in ERH frameworks. Parents described their sense of responsibility as immediate and instinctual, preceding an emotional bond. Parents demonstrated their agency in deciding what to carry forward from their relational histories, a pattern this study terms relational legacy. Integrating parent-generated language into ERH measurement research may shape a more comprehensive picture of ERH reflecting how families experience connection.

## INTRODUCTION

It is well established that a predominant factor for a child’s development and flourishing is an enduring emotional tie to at least one adult.^1-3^ When this parent-child relationship is of high quality, it enables the child to thrive socially, cognitively, and predicts better mental health outcomes throughout their life.^4-8^ In 2021, the American Academy of Pediatrics (AAP) published a paradigm-shifting statement endorsing a strong clinical focus on the positive elements of those parent-child relationships.^9^ This statement was in response to evidence of resilience building and buffering effects of close healthy relationships on adversity.^10^

In this statement, the AAP coined the term Early Relational Health (ERH), which they broadly described as safe, stable and nurturing relationships early in life.^9^ ERH houses several constructs ranging from more enduring features of the parent-child relationship, including attachment and bonding,^11^ to more proximal, moment-to-moment dimensions of the relationship, including parental sensitivity^12^ and parent-child emotional connection.^13^

Most theoretical constructs underlying ERH have been the product of research observations. Attachment theory, for example, was derived from Bowlby’s and Ainsworth’s observations of human and non-human primate behavior.^14^ There is a dearth of studies that qualitatively explore parents’ perspectives on developing a relationship with their children with the goal of identifying ERH constructs that are parent-centered and meaningful in the context of the daily lives of families. Of the few studies that qualitatively consider parents’ experiences of their early relationship with their child, even fewer consider the perspectives of mothers and fathers.^15^ Most use clinical samples, including mothers of preterm infants,^16^ parents of children with autism spectrum disorder,^17^ parents with trauma,^18^ or mothers who are at risk of losing custody of their children.^19^ When the sample is more normative, questions about the relationship are not central to the study focus.^20^ Without considering parents’ perspectives, ERH frameworks lack the conceptual rigor needed to guide effective clinical practice.

In 2022, in response to the AAP’s appeal to integrate ERH observations and interventions in pediatric primary care, the Center for Early Relational Health (CERH), Reach Out and Read (ROR) and Nurture Connection (NC) partnered to co-designed a parent-centered research agenda that 1-iteratively combines data-driven and practice-informed methodologies to generate foundational knowledge about strong ERH, and 2-develop strategies for real-world promotion of ERH in the pediatric primary care setting.^21^ As one early actionable step in this partnership, here we co-designed a qualitative study that foregrounds the perspectives of parents in its study design, data collection, and presentation of results with the goal of generating a parent-centered framework of ERH.

## METHODS

*Study Design and Community Partnership* A Community-Based Participatory Research (CBPR) framework, centering parent expertise across all phases of the research process was used.^22^ Parent expertise was ensured through six parent leaders who have worked on research projects previously with ROR. Parent leaders represented six communities of parents in Michigan, New Jersey, New Mexico, North Carolina, Pennsylvania, and Washington, including those who identify as Black, African American, or Brown; Indigenous parents; parents of children with special health care needs or disabilities; Spanish-speaking immigrant parents; parents with a Southern cultural background; and fathers.^23^ Parent leaders co-designed the interview guide, conducted interviews with parents from their networks, de-identified transcripts, and consulted twice at the data analysis stage. They also provided feedback on the naming of organizing principles to ensure language is parent-generated.

From July through December 2024, CERH/ROR researchers and parent leaders met five times to co-design a semi-structured interview guide, the “Parent-Child Connection Interview” (Table 1; Supplemental Files 1 & 2). Drawing on the Adult Attachment Interview^22^, the Parent Development Interview,^24^ the Working Model of the Child Interview,^25^ and the concept of emotional connection, the guide contained four sections: their own childhood relationships with their primary caregivers, current adult relationships, their relationship with their youngest child (0-4 years), and a brief demographic close. In the third section, parents told the story of their relationship, described their felt sense of connection, and recounted moments of feeling close or not close to the child. This nested structure reflected the study’s theoretical interest in situating moment-to-moment connection within both the overall parent-child relationship and the parent’s broader relational history.

**Table 1.** Example Questions from the Parent-Child Connection Interview.

| **Main Questions** | **Probes** |
| --- | --- |
| Please describe your relationships with your parents or primary caregivers (meaning the one or two people that took care of you most of the time when you were very young), starting from as early as you can remember. | • Were you able to go to them when you needed them?<br>• Were they there for you when you were sad or needed comfort?<br>• How did your relationship change over time? |
| Describe your current relationships with other adults. That can be family members, friends, colleagues, romantic partners. | • What are things that make you feel close to someone? It could be anything.<br>• Of the people you mentioned, is there one or more who you feel really close to? Tell me more why. |
| Tell me the story of your relationship [with your youngest child], starting from whenever you felt that it began. | • Did that feel like instant-falling in love or did the connection slowly form?<br>• Is that connection constant? Can you talk a little bit about how it changes over time? |
| Think of a recent moment that made you feel really close [to this child]. Describe it. | What made you feel close? What does it feel like when you feel really close? |
| Think of a recent moment where you didn't feel very close, or didn't feel like the best parent. Describe it. | What made you feel not-close? What does it feel like when you feel not-close? What happened after? |
| Can you tell me about yourself and your identity, or any identities that are important to you? | ...Whatever you're comfortable sharing. Examples are: if you identify with a particular culture, race, or ethnicity (can be more than one), or want to share about observing a religion or faith, or anything about your gender identity or sexual orientation. |

### Participants and Recruitment

Eighteen parents were recruited through six parent leaders’ personal and professional networks via snowball sampling. Recruitment was managed by parent leaders, a deliberate choice intended to reduce social desirability bias and facilitate candid sharing.

### Data Collection

Parent leaders had prior experience conducting semi-structured interviews. Prior to interviews, a CERH researcher provided a pre-recorded model interview and conducted a one-hour training addressing study-specific protocols (video-off Zoom recording, AI-assisted transcription via Vook.AI, and de-identification procedures) and best practices.^26^

Between May and August 2025, parent leaders each conducted one to four 60 to 90 minutes interviews (n=18 total). Parent leaders were free to adapt the interview guide as they saw fit. The peer-interviewing model was a deliberate methodological choice to support open sharing between parents.^27^ Demographic data were collected at the end of the interview. Five participants identified as fathers and 13 as mothers. The sample included participants who self-identified as White, Pacific Islander, American Indian, Black/African American, Native Hawaiian, or two or more of these categories. Most of our sample did not identify as Hispanic. The greatest number of participants lived in Pennsylvania and North Carolina, although participants from New Jersey, New Mexico, Washington, and Florida were included. (Due to small sample, sample sizes per category are not provided to preserve anonymity). Demographic data are included in Table 2.

**Table 2.** Demographic Characteristics of Participants.

| **Characteristic** | **Participants, No. (%) n=18** |
| --- | --- |
| Parent age, yr, mean (range) | 34.8 (29-45) |
| Parent sex |  |
| Female | 13 (72.2) |
| Male | 5 (27.8) |
| Marital status |  |
| Partnered/Married | 13 (72.2) |
| Single | 5 (27.8) |
| Number of children, mean, (range) | 2.7 (1-10) |
| Child age, months, mean (range) | 28.1 (9-48) |
| Child sex |  |
| Female | 8 (44.4) |
| Male | 10 (55.6) |
| Number of children in household |  |
| First child | 6 (33.3) |
| >1 child | 12 (66.7) |

Parent leaders also transcribed and de-identified all recordings prior to data being analyzed for research, maintaining community ownership of the data. Therefore, this study was determined to be Not Human Subjects Research by the Columbia University Institutional Review Board.

Sample size was guided by the principle of information power^28^ rather than data saturation, consistent with reflexive thematic analysis, which conceptualizes themes as generated through researcher engagement rather than as fixed entities to be exhaustively identified.^29^ The study’s focused aim, highly specific sample, and analytic strategy emphasizing interpretive depth supported the sufficiency of 18 interviews.

### Data Analysis

#### Analytic Framework

We used Braun and Clarke’s Reflexive Thematic Analysis (RTA)^29-31^ for its emphasis on conceptual depth and researcher transparency. In RTA, themes are patterns of shared meaning organized around a central organizing concept, forming an interpretive narrative rather than topic summaries. These principles were generated through a six-step process adapted from Braun and Clarke.^30^ Dedoose (Version 9.2.27)^32^ was used to organize analytic memos and track recurring patterns across transcripts.

#### Positionality

Reflexivity was central to our analytic process.^31^ The four CERH-based coders (K.F., M.W., S.D., D.M.) are all women under the age of 30 with professional experience as ERH researchers, and none are parents. Our team’s racial and ethnic backgrounds include three who identify as White and one as Hispanic/Latina. By contrast, the parent leaders’ demographic makeup closely mirrored participant diversity, making their involvement a critical corrective to the interpretive limitations of the research team. Positionality shaped coding in both generative and limiting ways: familiarity with ERH constructs supported recognition of theoretical significance in parent accounts, while pre-formed frameworks risked mapping experience onto existing constructs rather than allowing new meaning to emerge. Member-checking sessions, described below, were designed explicitly to counteract this latter risk.^33^

#### Six-Step Analytic Process

The team completed an initial pass through Steps 1 to 3 of the RTA framework.^30^ Coders read excerpts from all 18 transcripts (Step 1: Familiarization), developed a first collaborative codebook applied in pairs to all transcripts (Step 2: Generating Initial Codes), and grouped codes into broader themes (Step 3: Generating Themes).

In Step 4 (Reviewing Themes), a member-checking session with parent leaders revealed significant limitations in the initial approach: coders had focused their analysis primarily on the portions of the interview that directly addressed parents’ relationships with their youngest child, setting aside the earlier sections that explored parents’ own backgrounds and formative experiences. Parent leaders made clear that this separation was analytically untenable. A parent’s relationship with their child cannot be understood separately from the parent’s personal identity. The initial organizing principles did not adequately capture the depth or meaning of what parents had shared.

In response, the analytic cycle was repeated with the full dataset, reconstructing the codebook through independent coding and group reconciliation. The revised organizing principles were refined through a second member-checking session and a structured questionnaire completed by parent leaders.

In Step 5 (Defining and Naming Themes), the team used member-checking and questionnaire feedback to define and name the final organizing principles, prioritizing parent-facing language over researcher-imposed constructs. In Step 6 (Writing Up), the team used this refined framework to write the analytic narrative.

### Quality and Rigor

Team-based coding across 21 meetings between 11-2025 and 03-2026 ensured interpretive depth, supported by systematic analytic memos and the two member-checking sessions with parent leaders. Reporting followed the Standards for Reporting Qualitative Research (SRQR) checklist.^34^

## RESULTS

Together, the 18 interviews constructed a narrative of how early parent-child connection develops, is shaped by context, and transforms the parent over time. We generated six themes, which were further refined in member-checking sessions. When parent leaders were asked to rank the relevance of each theme, each leader selected a different theme, suggesting that all themes were important to this narrative.

The six themes are as follows: (1) Parent-child connection begins with instinctual responsibility. (2) From this foundation, connection builds as a dynamic process that ebbs and flows. (3) This early connection does not exist in isolation; it is directly influenced by the family environment. (4) Additionally, parents’ own experiences, upbringing, and past relationships indirectly shape how they connect with their child. (5) For connection to grow, parents must navigate internal and external influences to show up physically and emotionally for their child. (6) Over time, this intentional engagement leads to a profound transformation where personal growth of the parent is a key outcome. Importantly, parent leaders contributed directly to the naming of these six organizing principles through the member-checking sessions. Table 3 includes additional quotes from interviews.

**Table 3.**
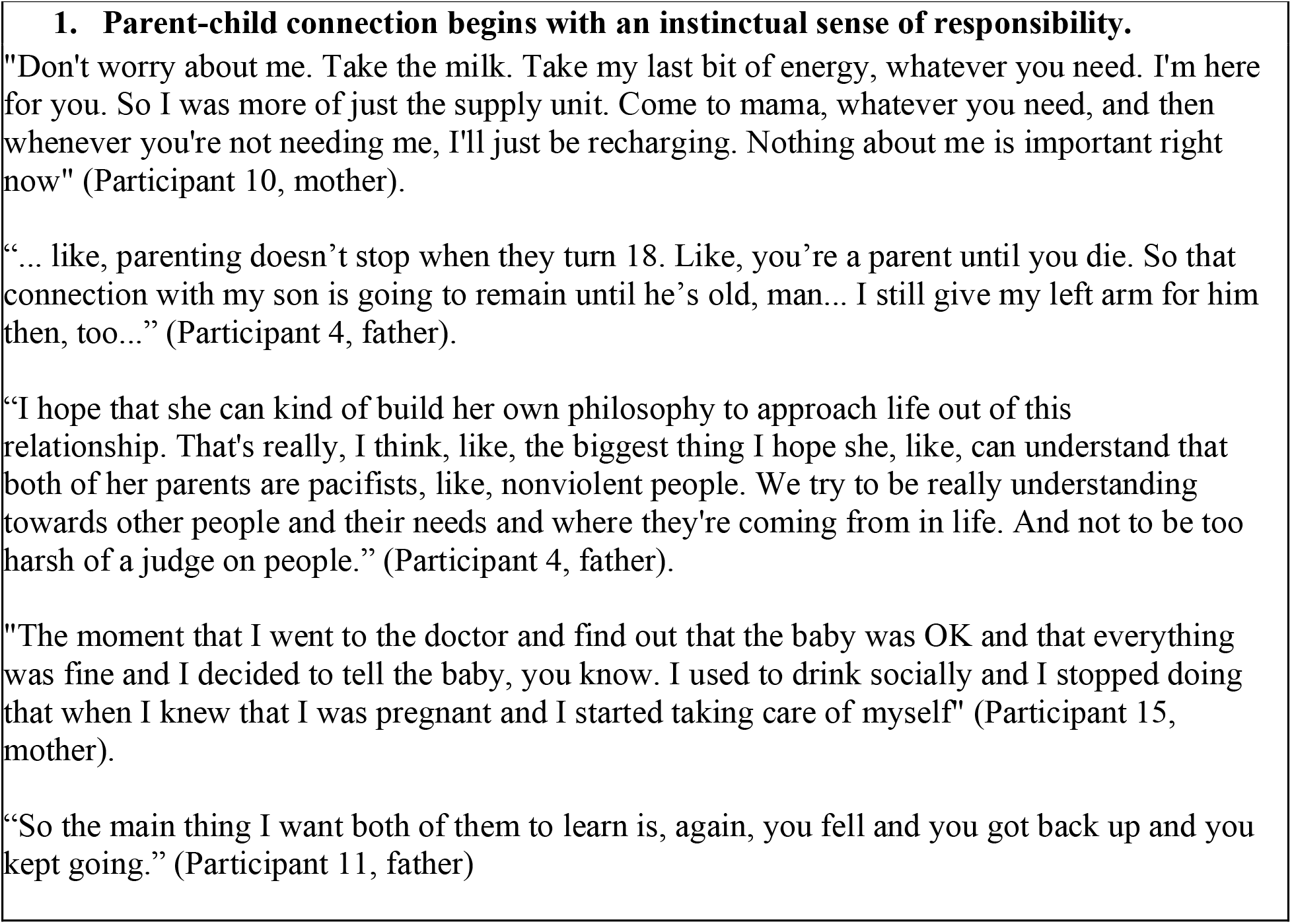
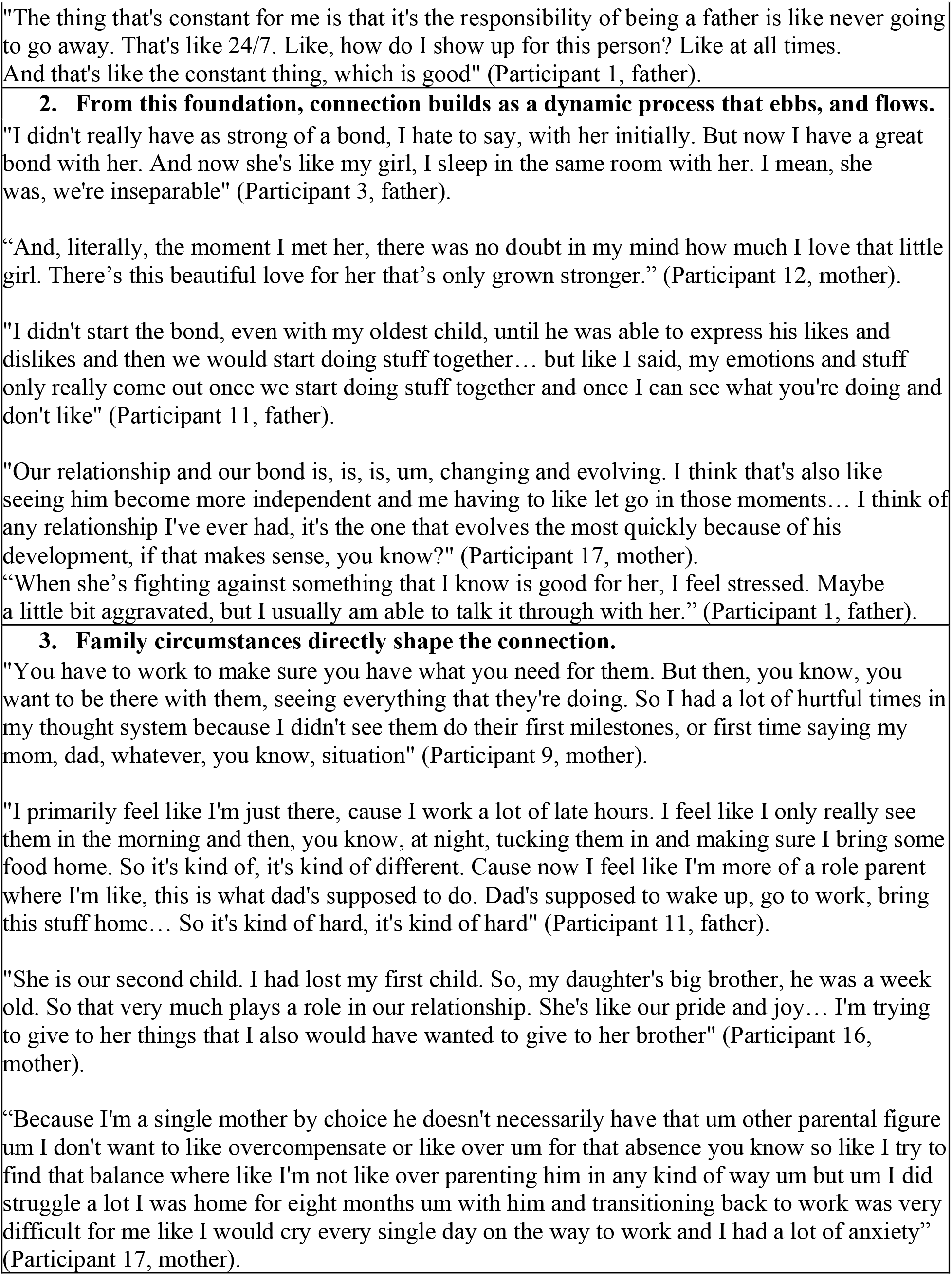
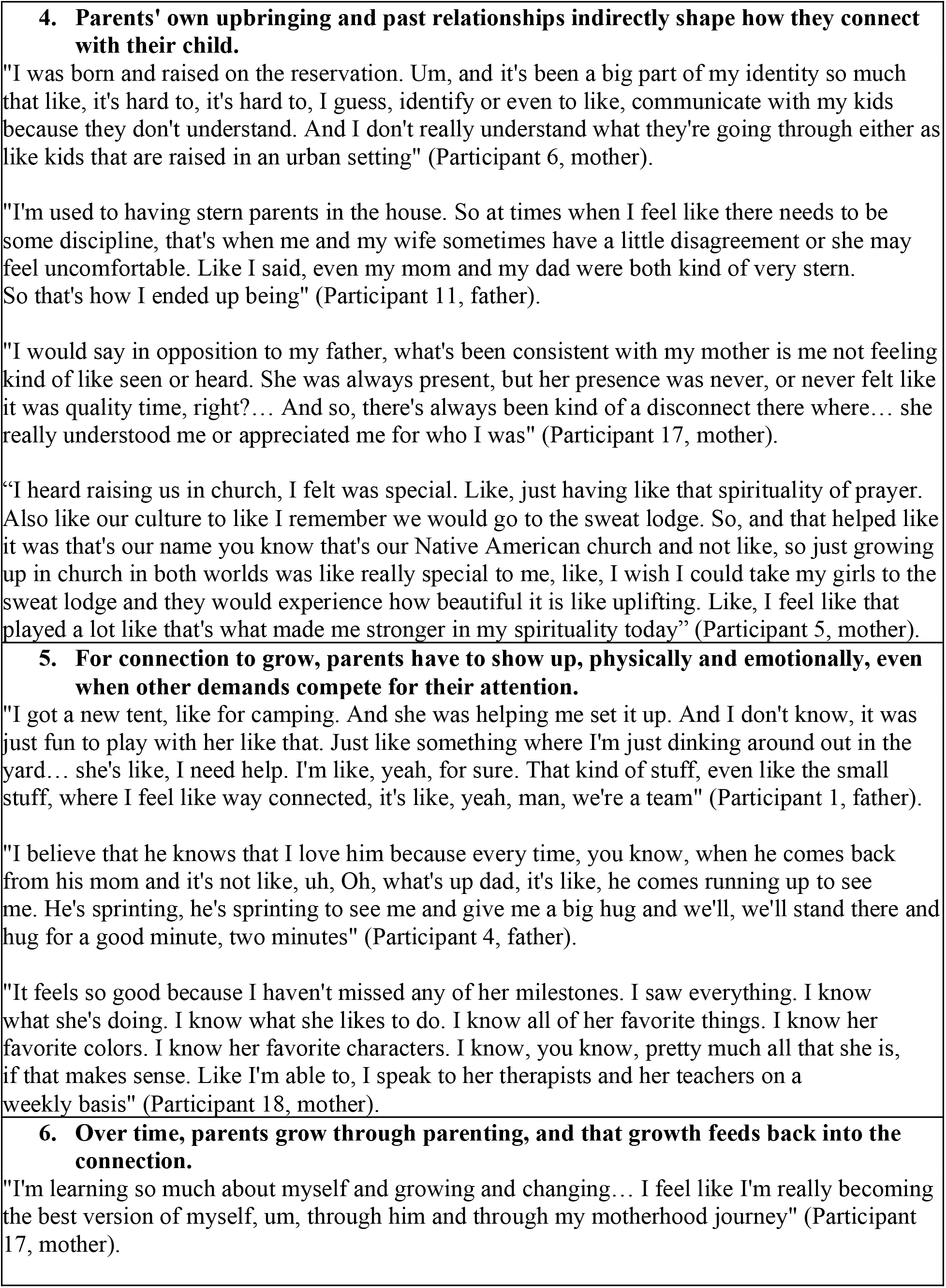
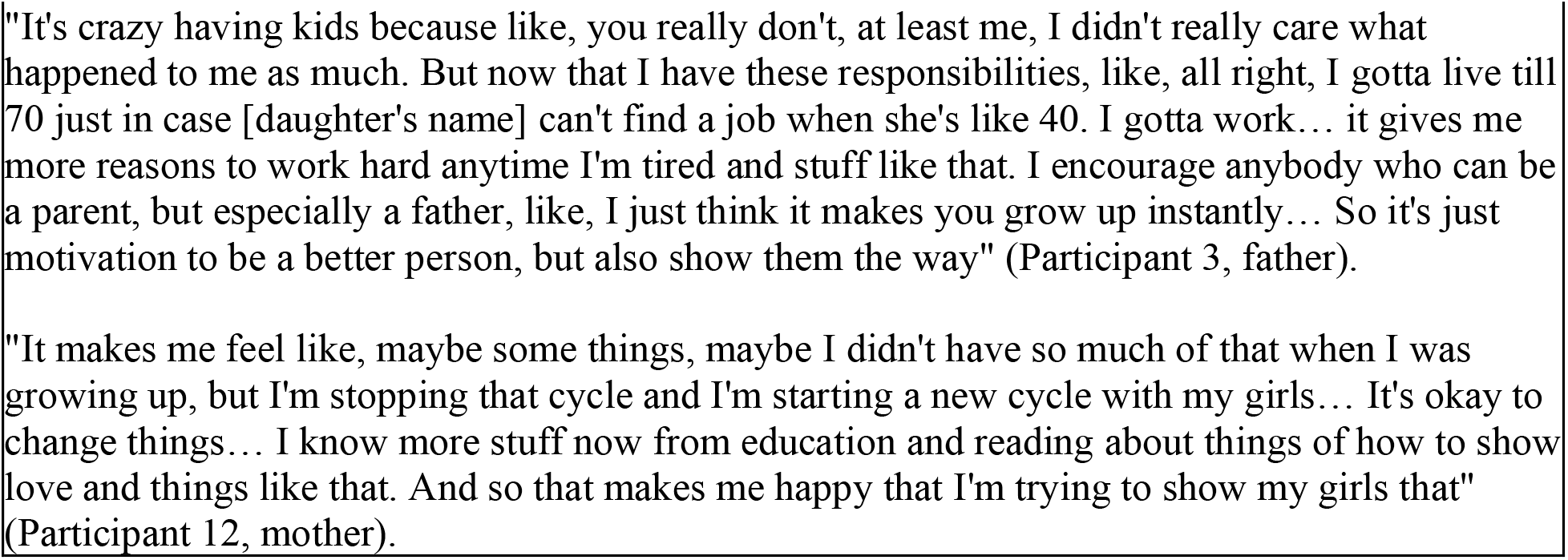
Mothers’ and Fathers’ Early Parent-Child Connection Quotations.

### 1) Parent-child connection begins with an instinctual sense of responsibility

In the interviews, parents described an internally motivated drive to care for their young children, establishing a baseline for safety and trust. Many parents shared that their sense of duty towards their child predated the development of the parent-child connection itself.

*“*… *I think love, you have to work on and build it. But it’s like, responsibility is, like, immediate. And, like, not just my responsibility, but just, like, I would put my own well-being below the well-being of this thing, this child that was just born*…*”* (Participant 2, father).

Additionally, several parents stated that this sense of instinctual responsibility persisted beyond when the child was born and represented a lifelong sense of responsibility for their children. Although the manifestation of that responsibility would likely evolve over time, such as transitioning from providing support with feeding and sleeping to more emotional support, the underlying sentiment of responsibility remained constant.

“*And I want them to know I’m always going to be there for them as best I can to push them along, to do their best*” (Participant 9, mother).

Building off of this enduring sense of responsibility towards their children, parents also expressed a sense of moral responsibility to instill values into their children.

“*It is my hope that he learns how to be his best advocate, because he sees and observes me do it on his behalf. But I hope that I’m not just doing that. I’m also advising him on how he can do it himself. Yeah, I like to be intentional when it comes to that with him*” (Participant 13, mother).

### 2) From this foundation, connection builds as a dynamic process that ebbs and flows

Connection is defined by constant adaptation by both the parent and child, which occurs through small, daily exchanges of care and communication such as eating or playing together. This connection develops dynamically, forming at different times relative to the child’s birth.

*“*… *I felt, like, my child’s presence in this bond for a very long time before he actually got here”* (Participant 17, mother).

*“So, most people would say, oh yeah, after my child was born, it was just instant. But for a long time, it felt like he was just exploring his world and navigating it to his own little drum, for sure. Or, and then, as the communication improved, I felt more connected to him in that way, right?”* (Participant 13, mother).

Moreover, parents described how the quality of connection changes over time. In other words, as the parent and child develop, so does the connection between them.

*“And it’s just like the stuff I was talking about with me and my dad, like it can ebb and flow over time based on how connected you’re staying and other circumstances in your life”* (Participant 2, father).

Finally, parents shared that their relationships with their children aren’t always constant. Moments of disconnection from their children were described as part of the parent-child relationship.

*“I remember there was, like, a time or two when she was first born that it was hard for me to look at her*… *So yeah, no, I struggled in the beginning”* (Participant 16, mother).

### 3) Family circumstances directly shape this connection

Parents emphasized that the early parent-child connection is strengthened or disrupted by external factors such as family structure, cultural expectations, and intergenerational values. These factors, such as birth order, the quality of the parents’ relationship with each other, and cultural norms, deeply influenced parenting choices and were described as inextricable from the development of parent-child connection. Some of these factors are experienced within the household, informing how connection manifests in day-to-day interactions.

*“My son’s mom has been very difficult to find that middle ground with, um, and that causes a big, uh, stress, but I try not to let it affect my time with my son and my relationship with my son. Cause we’re interacting in front of my son”* (Participant 4, father).

Others are broader and more abstract, such as the desire to break harmful intergenerational cycles by parenting their own children differently than they were parented.

*“And to see, the, how harsh my grandmother was, it was just like, oh gosh, my mom is so much nicer compared to her, but I think in order to reduce it, it’s never, it takes lifetimes*… *and I’m hoping for my boys and still my girls, you know, to see the flourishing of what I had been trying to reduce and what my mom had been trying to reduce and then break down”* (Participant 7, mother).

### 4) Parents’ own upbringing and past relationships indirectly shape how they connect with their child

During member-checking sessions, parent leaders emphasized the importance of recognizing that parents’ personal histories shape the context in which the parent-child relationship develops. Whereas the previous organizing principle highlights parents’ deliberate, conscious decision-making in response to external factors, this organizing principle centers on experiences that parents do not explicitly link to their relationship with their child. Although parents did not directly articulate the influence of these personal histories during interviews, we interpreted these experiences as exerting an indirect effect on the parent-child relationship, warranting careful consideration.

*“My parents were in prison. So, my dad went out when I was nine, six, six months. And my, my mom, when I think when I was around two, two or three. She went for a few years. And she got out and when she got out, she was in, like, rehab, and she came around, but she was just, like, a different person, like, a different, distant, mom*…*”* (Participant 5, mother).

*“I remember the one time that I did that I gave my mom the chance to step up and be a mom, and I let her know… that I was upset, and instead of trying to reach out and help, she just screamed at me, cussed me out, called me horrible names. So no, even to this day, I cannot rely on my parents for anything. I never could”* (Participant 14, mother).

### 5) For connection to grow, parents have to show up physically and emotionally, even when other demands compete for their attention

Regardless of external factors, clear patterns were evident in how early parent-child connection manifested in practice. For early parent-child connection to grow, active participation was required on the part of the parent. Parents must perform the work of being present even amidst the stressors and historical influences described in the preceding principles. Parents described this presence as two distinct domains.

The first was mutual emotional engagement. *“It’s like the joy I get from them is the same joy they get from me. And then what you put into your babies is what’s going to come out of them”* (Participant 9, mother).

The second was physical engagement. *“If there’s anything that child loves to do, we will just go outside and we will just roam. We’ll just walk around and have the best time in the world just walking. Yeah, she’s picking up rocks. Oh my God, my favorite thing”* (Participant 14, mother).

### 6) Over time, parents grow through parenting and that growth feeds back into the connection

This growth is informed by the parent’s relationship with their child as well as their experiences with their own caregivers, prompting them to better themselves to ensure that their children have better experiences than they did. This growth then comes full circle, directly informing and improving how the parent shows up for their child and thereby creating a self-sustaining cycle of relational health.

*“Man, I tell [child’s name] a lot, I’m just so grateful that she’s in my life. She reminds me to slow it down for her a bit, reminds me what it’s like to be a child again, which is […] weird because in a lot of way it feels like it heals a lot of childhood stuff for me, not that it’s [*…*] her burden, but she just does”* (Participant 1, father).

*“I’ve never struggled with anger or impatience ever. [*…*] And all of a sudden, now I’m here, I’m like, okay, like, I do have impatience in me. I do have anger in me. Um, I do have triggers that I didn’t know about, where like, I’m like [*…*] I will just have a really hard time staying emotionally regulated”* (Participant 8, mother).

## DISCUSSION

### Overview of Findings

Using peer-interviewing in which parent leaders recruited and interviewed participants from their own networks, six organizing principles were generated. Together, they portray early parent-child connection as a dynamic process: rooted in instinctual responsibility, shaped by the family environment and the parent’s own history, sustained through ongoing engagement, and ultimately a catalyst for the parent’s own growth, which feeds back to strengthen the relationship. Two principles pointing to gaps in current ERH conceptualization warrant dedicated discussion: instinctual sense of responsibility (Principle 1) and personal growth that feeds back into the relationship (Principle 6).

### Responsibility as an Understudied ERH Construct

Most current ERH frameworks rest on attachment theory and parental sensitivity,^2,35^ which emphasize how parental responsiveness to infant cues fosters security. What they do not explicitly address is responsibility as a discrete, internally motivated construct, which parents here described as foundational to their connection with their child. Parents described responsibility not as an externally imposed obligation but as an immediate, instinctual reorientation of priority: another person’s needs suddenly live alongside or ahead of their own. In their accounts, this shift was sudden yet enduring, present before love or emotional connection had fully formed, and endured even if its expression changed over time. During member checking, parent leaders affirmed that “instinctual sense of responsibility” best captured the essence of the construct. The construct is intrinsically different from sensitivity or bonding and may represent one of the earliest, most immediate expression of the broader psychological reorganization Stern^36^ termed the “motherhood constellation.” This reorientation of self both precedes and enables connection. Mothers and fathers both prioritized the child’s needs, though fathers more often framed this as duty and protection, and mothers as an embodied giving of self. With only five fathers, this difference is hypothesis-generating and warrants dedicated study.

### “Relational Legacy”: Extending the Intergenerational Transmission Framework

The intergenerational transmission of attachment is among the most studied concept in developmental science. Beginning with van IJzendoorn et al.’s^37^ meta-analysis, research has shown that parents’ attachment representations predict child attachment classification, and Verhage et al.^38^ confirmed these associations while identifying a persistent “transmission gap” that parental sensitivity alone does not explain. Fonagy et al.^39^ proposed reflective functioning, the capacity to understand behavior in terms of mental states, as one mechanism through which parents’ representations shape caregiving. Waters and Waters^40^ described attachment scripts as blueprints carried forward from early experience.

These frameworks are powerful but largely mechanistic: they describe how relational patterns transmit passively across generations, not how parents actively shape that transmission. Our data suggest something that is currently left underspecified. These parents were aware their history was present and were making deliberate choices about which patterns to carry forward and which to interrupt, i.e., not passively repeating their past. We propose the term *relational legacy* as an novel construct capturing three dimensions: 1-the inherited dimension (patterns, values, and experiences passed down across generations); 2-the parent’s deliberate reflection in deciding which to carry forward and which to interrupt; 3-the possibility of positive cycling, in which parents use the relationship itself as a site of healing and growth. While reflective functioning^39^ may be the mechanism that enables this deliberate selection, *relational legacy* is broader, spanning the inherited patterns themselves and their potential for positive cycling. This third dimension extends Fraiberg et al.’s^41^ “ghosts in the nursery” and Lieberman et al.’s^42^ “angels in the nursery”, where those frameworks centered, respectively, the intrusiveness of unresolved trauma and the protection of benevolent memories. *Relational legacy* encompasses the full process and centers parental agency over passive repetition. Crucially, relational legacy is not limited to attachment security; it spans the full multi-faceted ERH landscape, consistent with calls to move ERH beyond exclusively attachment-centric models.

### Implications

The organizing principles are meant to be hypothesis-generating. Connection as a dynamic, ebb-and-flow process calls for longitudinal, repeated-measures designs that capture within-dyad variability rather than cross-sectional snapshots. The family environment as shaping the connection calls for studies that model structural and cultural context, including family structures, especially those underrepresented in research.

We generated the principles of responsibility and personal growth from parents’ accounts, illustrating how CBPR can highlight dimensions of ERH that researcher-centric designs may have systematically overlooked. Responsibility and relational legacy merit particular attention. Responsibility calls for formal construct development and testing as a distinct predictor of ERH outcomes; because it often precedes bonding, it may also offer a low-stakes entry point for ERH conversations in well-child visits. Relational legacy could serve as a brief, strengths-based prompt: “What do you most want to carry forward from how you were raised? What do you want to do differently?” which could fit within a well-child visits and complement existing observation tools (e.g., ASQ:SE-2^43^ and EPDS).^44^

### Limitations

This was a qualitative study of 18 participants meant to generate hypotheses rather than test them. Recruitment through parent leaders’ networks may have introduced selection bias, and pre-existing relationships could have influenced responses. Participants were largely U.S.-born and engaged in ERH networks, which likely shaped how they articulated their experiences. Replication in broader community samples is needed, including cultural variation in how responsibility and intergenerational patterns are expressed.

## Data Availability

All data produced in the present study are available upon reasonable request to the corresponding author.

## Notes

### Competing Interest Statement

The authors have declared no competing interest.

### Author Declarations

IRB of Columbia University waived ethical approval for this work. On 9/25/25, the Columbia University IRB deemed the study (IRB-AAAV9559 (Y01M00)) "Not Human Subjects Research Under 45 CFR 46." The correspondence was as follows: "This protocol was reviewed by a member of the Human Research Protection Office (HRPO) Administrative Review Committee (ARC). The proposal does not meet the criteria to be considered human subjects research, as there is no interaction with subjects, there is no intervention, and private, identifiable information is not being collected. Thus, further IRB review is not needed."

### Funding

- Einhorn Collaborative
- Eunice Kennedy Shriver National Institute of Child Health and Human Development: K99HD115784

