## Supplemental Files 1 & 2 for "A Parent-Generated Framework of Early Connection: Findings from a CBPR Qualitative Study"

**Supplemental File 1. Final Draft: Parent-Child Connection Interview (02-2025)**
Note: Bolded questions are essential. Non-bolded questions are there to provide ideas of questions to ask to encourage the parent to delve deeper into their experiences.

### Introduction

1. Thank parent (in your own words) for being willing to meet with you.
2. Explain purpose: We are doing these interviews to learn about the different ways that parents and caregivers connect with their children, and how families and communities can be better supported for strong connections to form. We want to learn from the lived experiences of parents like you.
3. Your name is not connected to this interview and everything you say will be anonymous. Also, just so you know, nothing you tell me in this interview can be reported to any agencies such as social services.
4. The interview is audio-recorded (no video) and all of your words will be written down, so that we really learn from your own words.
5. We're open to every and all experiences. This is a judgment-free zone and anything you would like to share is welcome. The interview is meant to take about an hour. Some questions may feel too personal. You can skip any you don't want to answer. You can ask me questions at any point.
6. Is it OK to proceed?

### 1. Childhood relationships

1. **Tell me a little bit about the household you grew up in. Who lived with you? Where did you live (country or state)?**
2. **Now, please describe your relationships with your parents or primary caregivers (that means the one or two people that took care of you most of the time when you were very young), starting from as early as you can remember.** Describe one relationship first, then I'll ask some questions about it. If you had two caregivers, we'll do the same for the second relationship. [Parent leader: after they have described the relationship in some detail, ask the two questions below. Then repeat this process for the second parent or caregiver].
    a. **Can you describe that relationship, whether it was good or bad?** [This question is very important. After they respond, ask any follow-up questions that come to mind].
    b. Were you able to go to them when you needed them? Were they there for you when you were sad or needed comfort?
    c. How did your relationship change over time?
    d. Is there anything you feel was special or very specific to this relationship?
3. Some people are closest to adults who are not their primary caregivers. Was that the case for you? If so, was the person you were closest to when you were very young? Why them in particular?

### 2. Adult relationships

1. **Describe your current relationships with other adults. That can be family members, friends, colleagues, romantic partners.** [Parent leader, you can spend a few minutes on this question. We want to know who is in their social circle and a little bit about these relationships. Ask any follow-up questions that come to mind and invite them to share as

much about these relationships as they would like. This question is important–we want to learn as much as we can about their current relationships].

2. What are things that make you feel close to someone? It could be anything.
3. Of the people you mentioned before, is there one or more who you feel really close to?
    a. Tell me more why.

**<u>3. Relationship with child</u>**

1. **First, tell me about your current household. Who lives in your household?**
    a. How many children do you have, either living with you or not?
    b. How old are they?

Now I want you to focus on your youngest child (if twins, just pick one at random). [Parent leader, feel free to substitute in the child's name in the following questions, if the parent offers their name].

2. **What were your feelings when you found out you were going to have this child?**
3. **Tell me the story of your relationship, starting from whenever you felt that it began.**
    a. **How connected do you feel to this child?**
        i. If connected,
            1. When did that connection start?
            2. Did that feel like instant-falling in love or did the connection slowly form?
            3. Do you think this was how it happened for your child?
            4. **Can you describe what connection with your child feels like to you?**
            5. Is that connection constant? Can you talk a little bit about how it changes over time?
            6. Think of a recent moment that made you feel really close. Describe this moment.
                a. What made you feel close?
                b. What does it feel like when you feel really close?
        ii. If not connected,
            1. **What does that feel like to you? To feel not so connected?**
            2. Have you always not felt very connected?
            3. Is that okay for you? Or do you wish you felt more connected to this child?
            4. Think of a recent moment where you didn't feel very close, or didn't feel like the best parent. Describe this moment.
                a. What made you feel not-close?
                b. What does it feel like when you feel not-close?
                c. What happened after? Did something make it better?
4. **If your relationship was a book, movie, TV show, or song, what would it be called?**
5. What are some things you and your child like to do together when you're interacting?
6. [If they have more than one child]: How is your relationship similar or different to those you have with your other children?
7. What do you hope your child learns from this relationship?
8. Now, please describe a typical day for your child.
    a. During a typical weekday, who cares for your child (it can be more than one person)?

b. How much time do you spend together on a typical day?
c. Do you wish you had more time or would you prefer it be less?

9. **What does screen time usually look like for you, not including work-hours?**
    a. **What about screen-time for your child?**
10. **Would you be comfortable sharing any family challenges or stressors?** It could be anything related to your children, household, health, or financial situation.
11. Is there anything else you would like to tell me about your relationship with your child?

**<u>4. Demographic information</u>**

1. **How old are you?**
2. **Where were you born (country or state)?**
3. **Where do you currently live (state)?**
    a. When did you move to the U.S. (if it applies)?
4. **Can you tell me about yourself and your identity, or any identities that are important to you?** Whatever you're comfortable sharing. Examples are: if you identify with a particular culture, race, or ethnicity (can be more than one), or want to share about observing a religion or faith, or anything about your gender identity or sexual orientation.

**<u>Ending interview</u>**

1. End interview with a heartfelt expression of gratitude (in your own style).
    a. Something like: Thank you so much for giving me your perspective, telling me about your experiences, and sharing your story with me.
2. Can ask: Do you have any questions about this interview or the purpose of this project?

**<u>Supplemental File 2. First Draft: Parent-Child Connection Interview (07-2024)</u>**

**<u>Introduction</u>**

Thank you so much for agreeing to do this interview. I'm going to ask you about the important relationships in your life. I want to learn about your relationships as a child, as an adult, and especially, how you view your relationship with your child.

You can share as much or little as you're comfortable sharing. Everything you share will remain confidential within the study team. Some questions might feel too personal, and you can skip any questions you aren't comfortable answering.

If you've agreed to have the interview recorded, it will be stored in a secure Medical Center server with no link to your name or other identifying information. It will be transcribed by trained researchers and use of anything you say will remain anonymous.

**<u>1. Demographic information</u>**

1. How old are you?
2. Where were you born (country or state)?
3. Where do you live now (state)?
    a. How many years have you lived in the U.S. (if applicable)?
4. What is your cultural background?
5. What is your race and/or ethnicity?
6. Are you a follower of a faith or religion? Which?

7. What's your highest level of education (high school, college, graduate school)?
8. Do you have active health insurance? Is it public or private?

**2. Childhood relationships**

1. Can you start by telling me about the household you grew up in–who you lived with, where you lived, if you moved around much, what your family members did at different times for a living?
    a. Would you say this was a very social household, always open to friends and visitors, or a more private, quiet household that kept to itself?
2. What would you say was most important to the people who raised you? Some examples could be supporting the family, going to church, helping your community.
3. What mattered to them in terms of how you should be? For example, was it important that you got good grades, were athletic, helped take care of your siblings?
    a. Did they raise you to behave in any specific ways in terms of how you interacted with others? For example, was it important that you had good manners, obeyed family rules, or expressed your own needs, feelings, and thoughts?
4. Describe your relationships with your parent(s)/caregiver(s) as a young child, starting from as far back as you can remember.[2]
    a. As a young child, which adult(s) were you closest to?
    b. Why would you say you were closest to this person/these people?
    c. When you were upset as a young child, who would you go to to be comforted?

**3. Relationships in adult life**

1. Can you describe to me what your friendship circle looks like?
    a. How many friends do you feel extremely close to, like you could share anything with?
    b. How comfortable are you depending on friends for emotional support?
    c. How many of your friends do you feel really know you?

2. Are you currently in a romantic relationship?
   If yes…
    a. What's your partner's name?
    b. In a few words, can you tell me about your relationship with [partner's name]–how you met, how long you've been together, how often you see [him/her/them]?
    c. Is [partner's name] the parent of your child/children?
    d. How do you feel about your relationship and how things are between you?
    e. When there's conflict, what does that look like? Is there yelling? Is one person a yeller and one quieter? Are both of you yellers? Both quiet?
    f. How does the conflict usually end?
    g. What would you say is the main purpose this relationship serves for you both?

   If no…
    h. Are you dating at all, or seeing anyone in a casual sense?
    i. How do you feel about this relationship-status–would you like to be in a romantic relationship or not?

j. In a few words, can you tell me about your last relationship–when it ended, how long you were together?

3. Now, I'd like you to think of a recent moment where you felt very close to and emotionally connected with someone you know pretty well (an adult). It could be a romantic partner, family-member, or close-friend.
    a. Describe this moment.
    b. What made you feel close to/connected with this person?
    c. How would you describe this feeling of closeness/connection with him/her/them?
    d. What does it mean to you to be emotionally connected with someone?

4. Now, I'd like you to think of a recent moment where you felt a bit emotionally disconnected from someone (an adult).
    a. Describe this moment.
    b. What made you feel disconnected from this person?
    c. How would you describe this feeling of disconnection with him/her/them?

5. Do you ever find yourself feeling lonely?
    a. Much of the time or only sometimes?
    b. Can you describe what <u>loneliness</u> feels like to you?

**<u>4. Relationship with child</u>**

1. Tell me about your child/children–how many children do you have, how old are they?

Please focus on your <u>youngest</u> child for the remaining questions.

2. What is this child's name?
3. In just a few sentences, describe a typical day for [child's name].
    a. How many hours a day do you spend playing together?
        i. Reading together?
    b. Is [he/she] in childcare?
    c. How many hours per day, not including work-hours, do <u>you</u> spend in front of a screen?
    d. How many hours per day does [child's name] spend in front of a screen?
4. What are the things that led up to the pregnancy? Was it planned or unplanned? How did you decide to have [child's name]?
5. Tell me about your experiences with the pregnancy and birth of [child's name].
    a. What were these experiences like for you, physically (if mother)? Emotionally?
6. Tell me about the first time you felt a connection with [child's name].
    a. How could you tell you felt connected?
    b. Was it easy or hard for you to feel connected with [child's name] in the very beginning?
    c. Were you ever worried you would not feel connected to [child's name]?
7. Has your relationship with [child's name] changed over time? How so?
8. If you could give your relationship with [child's name] a title, as if it were a book or movie, what would it be?

9. Think about a recent moment of connection with [child’s name] that made you feel really good.
    a. Describe this moment.
    b. What made you feel connected?
    c. Can you describe what connection with [child’s name] feels like to you?
10. Think about a recent moment of disconnection with [child’s name] that made you feel not so good.
    a. Describe this moment.
    b. What made you feel disconnected?
    c. Can you describe what disconnection with [child’s name] feels like to you?
11. If you could change one thing about your relationship with [child’s name], what would that be?
12. If there is one thing you would say is special about your relationship with [child’s name], what would that be?
13. If you could go into the mind of [child’s name], how do you think he/she sees you?
14. If there is one thing about your relationship that you hope every other parent and child could also experience, what would that be?
15. If I were to meet [child’s name] 20 years from now, what would you hope that he/she tells me about your relationship?

**Closing remarks**

Those were all of the questions I had for you. Is there anything else you think would be helpful for me to know about your relationship with [child’s name] or other relationships?

Thank you for your time. It’s been so helpful for me to hear about your experiences, and this interview will help us help parents to build stronger, more joyful relationships with their children from an earlier age, since it’s these connections that allow us to thrive in the world.
